# Prediction of Preeclampsia from Clinical and Genetic Risk Factors in Early and Late Pregnancy Using Machine Learning and Polygenic Risk Scores

**DOI:** 10.1101/2023.02.03.23285385

**Authors:** Vesela P Kovacheva, Braden W Eberhard, Raphael Y Cohen, Matthew Maher, Richa Saxena, Kathryn J Gray

**Affiliations:** Department of Anesthesiology, Perioperative and Pain Medicine, Brigham and Women’s Hospital, Harvard Medical School, Boston, MA; PathAI, Boston, MA; Center for Genomic Medicine, Massachusetts General Hospital, Boston, MA; Department of Anesthesia, Critical Care and Pain Medicine, Massachusetts General Hospital, Harvard Medical School, Boston, MA; Division of Maternal-Fetal Medicine, Brigham and Women’s Hospital, Harvard Medical School, Boston, MA

**Keywords:** preeclampsia, pregnancy, blood pressure, polygenic risk scores, machine learning, risk factors

## Abstract

**Background:** Preeclampsia, a pregnancy-specific condition associated with new-onset hypertension after 20 weeks gestation, is a leading cause of maternal and neonatal morbidity and mortality. Predictive tools to understand which individuals are most at risk are needed.

**Methods:** We identified a cohort of N=1,125 pregnant individuals who delivered between 05/2015-05/2022 at Mass General Brigham hospitals with available electronic health record (EHR) data and linked genetic data. Using clinical EHR data and systolic blood pressure polygenic risk scores (SBP PRS) derived from a large genome-wide association study, we developed machine learning (xgboost) and linear regression models to predict preeclampsia risk.

**Results:** Pregnant individuals with an SBP PRS in the top quartile had higher blood pressures throughout pregnancy compared to patients within the lowest quartile SBP PRS. In the first trimester, the most predictive model was xgboost, with an area under the curve (AUC) of 0.73. Adding the SBP PRS to the models improved the performance only of the linear regression model from AUC 0.70 to 0.71; the predictive power of other models remained unchanged. In late pregnancy, with data obtained up to the delivery admission, the best performing model was xgboost using clinical variables, which achieved an AUC of 0.91.

**Conclusions:** Integrating clinical and genetic factors into predictive models can inform personalized preeclampsia risk and achieve higher predictive power than the current practice. In the future, personalized tools can be implemented in clinical practice to identify high-risk patients for preventative therapies and timely intervention to improve adverse maternal and neonatal outcomes.

## INTRODUCTION

Preeclampsia, defined as new onset of elevated blood pressure after 20 weeks gestation, is a leading cause of maternal and neonatal morbidity and mortality worldwide.^1^ Preeclampsia affects 2-8% of all pregnancies^2^ and contributes to 26% of maternal deaths worldwide and 15% of preterm births^3^. In the US, preeclampsia incidence is increasing and results in significant healthcare utilization.^2^ Maternal complications include end-organ damage, eclamptic seizures, and death. Fetal/neonatal complications include growth restriction and iatrogenic preterm birth. Timely diagnosis and treatment can reduce the risk for severe maternal and neonatal morbidity by 72-89%.^4,5^

Current clinical practice in patients at risk for preeclampsia is focused on close surveillance, early detection, and prompt management.^6,7^ Pregnant patients’ risk for preeclampsia is assessed at the first prenatal visit and, in those at high risk, prophylaxis with low-dose aspirin and close blood pressure monitoring is recommended. Patients at high risk for preeclampsia should be managed by providers with experience in high-risk pregnancy at tertiary care hospitals. Currently, high-risk individuals are identified based on clinical factors, including pre-existing hypertension, obesity, pregestational diabetes, advanced maternal age, multiple gestation, and prior preeclampsia; however, this approach fails to identify 46-60% of pregnancies that develop preeclampsia.^8-10^ Improved tools to understand each individual’s personalized disease risk has the potential to markedly improve pregnancy care and clinical outcomes. Machine learning methods, based on implicitly learning relationships in large datasets allow for precise outcome prognostication and may improve preeclampsia prediction. Recent machine learning studies about the risks of hypertensive disorders of pregnancy^11^ and preeclampsia^10,12^ demonstrate the potential of these methods to generate highly accurate predictions.^13^ However, models published to date have low predictive power in early pregnancy when little clinical information is available; in addition, a significant number of patients – especially nulliparous patients without clinical risk factors – develop preeclampsia and, thus, fail to be identified by current models.

While preeclampsia has substantial heritability based both on maternal and fetal factors, ^14^ the specific genetic factors contributing to risk are just beginning to be identified, as detailed in recent genome-wide association studies (GWAS).^15-17^ Importantly, in the largest published maternal preeclampsia GWAS, the top hits were all loci previously implicated in essential hypertension risk. In addition, several studies have demonstrated that the overall genetic architecture of maternal preeclampsia overlaps with the genetics of both systolic and diastolic blood pressure, as well as body mass index (BMI).^15-18^ Given that essential hypertension is a known clinical risk factor for preeclampsia,^19,20^ and genetic predisposition to hypertension is associated with increased preeclampsia risk, we hypothesized that a machine learning model incorporating both clinical risk factors and a hypertension genetic risk score (i.e., polygenic risk score, PRS, generated from GWAS summary statistics^21,22^) could improve preeclampsia risk prediction for pregnant individuals. As PRS are associated with disease risk independent of other clinical and environmental risk factors, all factors can be combined additively in a single model. ^23,24,25^

In this study, we utilize a rich database derived from the electronic health record (EHR) of patients who have had a pregnancy in our healthcare system linked with genetic data from the biobank. We investigate the relative importance of different clinical risk factors and polygenic risk scores in the first trimester, as well as late pregnancy (before admission for delivery), to predict preeclampsia.

## METHODS

The data that support the findings of this study are available from the corresponding author upon reasonable request and based on institutional guidelines.

### Population

This study was approved by the Mass General Brigham Institutional Review Board, protocol # 2020P002859, with a waiver of patient consent. Pregnant patients were selected based on documentation of pregnancy greater than 20 weeks gestation and associated billing codes for cesarean or vaginal delivery. We included all available patients from May 2015 to May 2022 with genetic data available in the Mass General Brigham Biobank and analyzed each pregnancy independently. These dates were chosen as May 2015 is when our institution implemented electronic health records across all outpatient offices and inpatient sites. All data (including sociodemographic, clinical diagnoses, laboratory, vital signs, and genotyping) was obtained and analyzed using our machine learning platform,^26^ which extracts, transforms, and harmonizes data from multiple sources. Preeclampsia diagnosis was based on the established American College of Gynecologists and Obstetricians guidelines.^6^ All preeclampsia cases (N =87) were further validated by an experienced clinician.

### Genotyping and Imputation

Genome-wide genotyping for each patient was obtained from the Mass General Brigham Biobank,^27^ a prospective biobank launched in 2010 that contains genotyping data, samples, and questionnaires with ongoing links to tEHR. This effort is continuing, with 129,000 patients enrolled and more than 56,000 genotyped. Genotyping was performed using one of two Illumina single nucleotide polymorphism (SNP) Arrays: the MultiEthnic Genotyping Array (containing >1.6M SNPs) or the Global Screening Array (containing > 575K SNPs). Imputation was performed using the TOPMed Imputation Server.

### Polygenic Risk Scores

As systolic blood pressure (SBP) is the trait with the highest genetic correlation with the genetics of maternal preeclampsia and has the highest predictive power for future hypertensive disorders and cardiovascular disease^29^, we created an SBP PRS using the open-source PRS-CS tool.^28^ PRS-CS computes SNP effect sizes by high-dimensional Bayesian regression using GWAS summary statistics and a linkage disequilibrium reference panel. We selected the largest blood pressure GWAS meta-analysis to date, with over 1 million individuals,^19^ and used a European linkage disequilibrium reference panel with 1.1 million variants derived from samples from the 1000 Genomes Project to create SBP PRS in our study population. We categorized the PRS into quartiles of risk ranging from lowest to highest genetic risk: <25%, 25-49%, 50-75%, and >75%. We adjusted all models in which the SBP PRS was used by the first 10 principal components of ancestry (PCAs) to account for population structure.

### Statistical Analyses and Definitions

For the analyses, we used all available EHR data from before conception to up to 6 weeks postpartum. Variables were treated as parametric or non-parametric according to their distribution; continuous parametric variables were expressed as mean ± SD, and nonparametric variables as the median with interquartile range (IQR). Significance was determined using the Student’s t-test and one-way ANOVA for parametric variables, the Kruskal-Wallis rank sum test for non-parametric variables, and Fisher’s exact or Chi-squared test for categorical variables. A p-value of less than 0.05 was considered significant.

### Machine Learning and Linear Regression Predictive Models

Our machine learning platform,^26^ which utilizes Python 3.9 (sci-kit learn library), was used for the development of predictive models. We selected established clinical risk factors known to be associated with preeclampsia risk in published studies and guidelines. ^6,7,9,10,12,30^ For the predictive models, we created datasets in which only data obtained up to the selected time point was included to minimize the risk of data leakage. When adding the SBP PRS to the models, we considered the PRS as an independent predictor and adjusted by the first 10 PCAs. When incorporating time-series data of blood pressure measurements across pregnancy, we divided the pregnancy period into the following intervals: 0-14, 14-20, 20-24, 24-28, 28-32, and >32 weeks gestation. When incorporating time-series data for pregnancy weight gain, we used the BMI measured in the following intervals: 0-14, 14-28, and >28 weeks gestation. To assess the discrimination performance of the models, receiver operator characteristic curves (ROC) were developed, and the area under the curve (AUC), accuracy, sensitivity, specificity, and precision were calculated.

## RESULTS

### Patient Characteristics

Of 105,673 pregnancies recorded in our healthcare system after May 2015, genotyping data were available for 1,125 pregnancies (828 unique patients), all of whom were included in the study. The patient population was multi-ancestry, with 32.7% of patients self-identifying as non-White. Of the 1,125 pregnancies, 87 had a clinical diagnosis of preeclampsia (7.8%). Patients with preeclampsia were older and more likely to be nulliparous (Table 1). Patients who self-identified as Black or Hispanic were more likely to have hypertension and were more likely to develop preeclampsia (p<0.01). In addition, patients with any hypertensive disorder, including preeclampsia, chronic, or gestational hypertension, were more likely to have a family history of chronic hypertension and preeclampsia compared to normotensive patients (p<0.01). Patients with preeclampsia delivered before the 37^th^ week of gestation more often as compared to patients who were normotensive or who had chronic or gestational hypertension. As expected, patients with preeclampsia had the highest systolic and diastolic blood pressure during pregnancy compared to those with chronic or gestational hypertension, and normotension (p<0.01).

**Table 1.** Pregnant patient clinical characteristics

|  | **Preeclampsia (n=87)** | **Chronic and gestational hypertension (n=95)** | **Normotensive (n=943)** | **P-value** |
| --- | --- | --- | --- | --- |
| Maternal age at delivery, y | 32.9 (29.5 - 36.4) | 34.4 (30.5 - 37.8) | 33.5 (30.5 - 36.3) | 0.27 |
| Self-reported race |  |  |  |  |
| White | 60 (69%) | 56 (67%) | 691 (72%) | 0.8 |
| Black | 13 (15%) | 14 (17%) | 61 (6%) | < 0.01 |
| Other | 15 (17%) | 15 (18%) | 211 (22%) | 0.49 |
| Self-reported ethnicity |  |  |  |  |
| Hispanic | 3 (3%) | 4 (5%) | 41 (4%) | 0.91 |
| Non-Hispanic | 84 (97%) | 80 (95%) | 913 (96%) | 1 |
| Hospital |  |  |  |  |
| Tertiary | 84 (97%) | 82 (98%) | 886 (93%) | 0.87 |
| Community | 3 (3%) | 2 (2%) | 68 (7%) | 0.13 |
| Gravidity | 2.0 (1.0 - 3.0) | 2.0 (2.0 - 4.0) | 2.0 (1.0 - 3.0) | 0.15 |
| Parity | 1.0 (0.2 - 2.0) | 1.0 (0.5 - 2.0) | 1.0 (1.0 - 2.0) | 0.35 |
| Gestational age at delivery, weeks | 37.1 (35.3 - 38.3) | 38.3 (37.0 - 39.1) | 39.3 (38.6 - 40.1) | < 0.01 |
| Last BMI before pregnancy, kg/m<sup>2</sup> | 29.3 (23.8 - 34.3) | 28.7 (24.5 - 34.3) | 24.9 (22.0 - 29.2) | < 0.01 |
| BMI at delivery (kg/m<sup>2</sup>) | 32.9 (28.6 - 37.86) | 33.1 (30.0 - 37.5) | 29.62 (26.3 - 33.2) | < 0.01 |
| Maximal SBP during pregnancy, mmHg | 151.0 (142.0 - 160.5) | 146.0 (135.5 - 154.0) | 128.0 (120.0 - 136.0) | < 0.01 |
| Maximal DBP during pregnancy, mmHg | 93.0 (87.2 - 98.7) | 91.0 (84.0 - 94.7) | 80.0 (74.0 - 84.0) | < 0.01 |
| Family history of chronic hypertension | 39 (45%) | 55 (65%) | 409 (43%) | 0.012 |
| Family history of preeclampsia | 1 (1%) | 4 (5%) | 11 (1%) | 0.028 |

Median (IQR) for continuous variables; n (%) for categorical variables; p-values for continuous variables based on Kruskal-Wallis rank sum test; for categorical variables based on Fisher's exact or Chi-squared test.
Abbreviations: SBP, systolic blood pressure; DBP, diastolic blood pressure, BMI, body mass index

### Polygenic Risk Scores and Maternal Blood Pressure

Patients with SBP PRS in the highest quartile had higher maximal systolic and diastolic blood pressure during pregnancy compared to patients with the lowest quartile SBP PRS (Table 2). Patients with higher SBP PRS were more likely to be diagnosed with a hypertensive disorder (preeclampsia, chronic or gestational hypertension); in contrast, patients with lower SBP PRS were more likely to be normotensive throughout gestation, p<0.05. As SBP PRS was developed using a European population, we performed a sensitivity analysis applying SBP PRS only in the subset of the population that self-identified as White. This sensitivity analysis demonstrated similar findings (Suppl. Fig. 1) and additionally identified that patients with the highest PRS had a higher incidence of chronic hypertension. Also, patients with any hypertension diagnosis (gestational, chronic, or preeclampsia) had higher SBP PRS compared to normotensive patients (Suppl. Fig. 2).

**Table 2.** Patient clinical characteristics by lowest and highest quartiles of SBP PRS

|  | PRS<25% (n=280) | PRS>75% (n=281) | p-value |
| --- | --- | --- | --- |
| Self-reported race |  |  |  |
| White | 134 (48%) | 255 (91%) | < 0.01 |
| Black | 74 (26%) | 2 (1%) | < 0.01 |
| Other | 74 (26%) | 26 (9%) | < 0.01 |
| Self-reported ethnicity |  |  |  |
| Hispanic | 19 (7%) | 2 (1%) | < 0.01 |
| Non-Hispanic | 261 (93%) | 279 (99%) | 0.46 |
| Chronic hypertension | 14 (5%) | 23 (8%) | 0.14 |
| Gestational hypertension | 3 (1%) | 12 (4%) | 0.021 |
| Preeclampsia | 22 (8%) | 24 (9%) | 0.78 |
| Any Hypertensive disorder | 39 | 59 | 0.04 |
| SBP at the first prenatal visit, mmHg | 119.0 (112.0 - 128.0) | 117.0 (110.0 - 125.7) | 0.029 |
| DBP at the first prenatal visit, mmHg | 73.0 (68.0 - 79.0) | 72.0 (66.0 - 80.0) | 0.71 |
| Maximal SBP at any time in pregnancy, mmHg | 140.0 (130.0 - 148.0) | 142.0 (133.0 - 156.0) | < 0.01 |
| Maximal DBP at any time in pregnancy, mmHg | 86.0 (80.0 - 92.0) | 89.0 (81.0 - 96.0) | < 0.01 |
| Antihypertensive drugs during pregnancy | 19 (7%) | 22 (8%) | 0.65 |
| Family history of chronic hypertension | 130 (46%) | 122 (43%) | 0.59 |
| Family history of preeclampsia | 5 (2%) | 6 (2%) | 0.77 |

Median (IQR) for continuous variables; n (%) for categorical variables; p-values for continuous
variables based on Kruskal-Wallis rank sum test; for categorical variables based on Fisher's
exact or Chi-squared test.
Abbreviations: SBP, systolic blood pressure; DBP, diastolic blood pressure, PRS polygenic risk scores

### Models to Predict Preeclampsia

We sought to predict patient preeclampsia risk at two-time points – early in pregnancy, at the first prenatal visit, and late in pregnancy, before admission for delivery. If a patient had a preeclampsia diagnosis or delivered before the time point, any data after that event was excluded to minimize data leakage. Because relationships between predictors may not be linear, we developed both linear regression and nonlinear machine learning models. Subsequently, we investigated if the addition of SBP PRS improved the predictive power of the respective model and evaluated the predictive power of each model using only clinical, only genetic, or both genetic and clinical variables, respectively (Table 3).

**Table 3.**
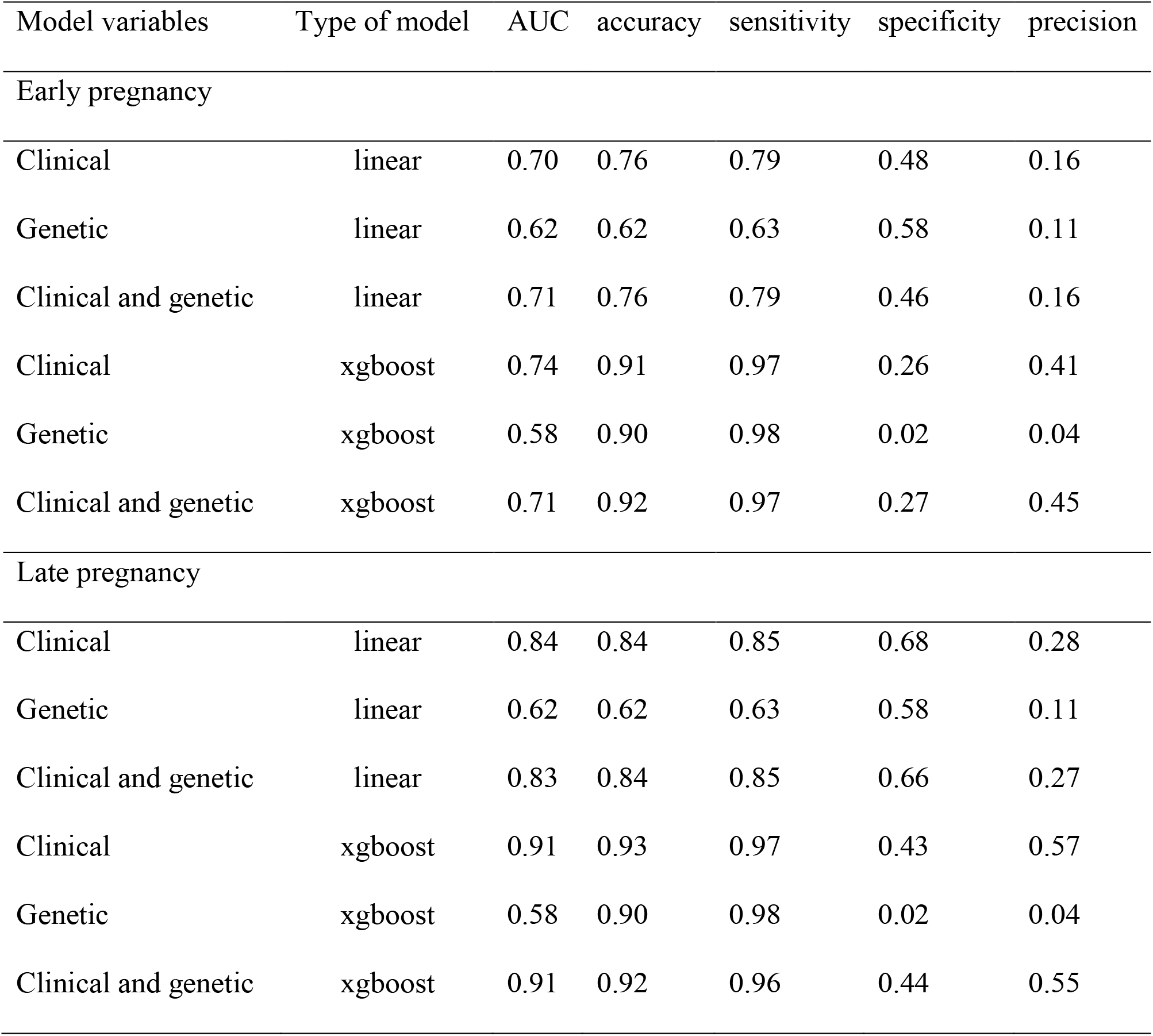
Power of clinical, genetic, and combined models to predict preeclampsia in the patient cohort using linear regression or machine learning models (n=1125)

In early pregnancy, patients are screened for preeclampsia risk based on the presence of established clinical risk factors. We used these risk factors to develop predictive models (Suppl. Table 1). The relationship between all variables is shown in Fig 1A. The clinical linear regression model, which was developed using only clinical variables available up to 14 gestational weeks, had an AUC of 0.70 (Table 3). We also created a separate genetic linear regression model using only SBP PRS, adjusted for the PCAs; this model had a weak predictive power, AUC 0.62. Adding the SBP PRS to the clinical risk factors in a combined linear regression model increased the AUC to 0.71, which was higher than either the clinical or PRS models alone. As machine learning allows for the incorporation of multiple variables with complex relationships, we developed a clinical xgboost model, which achieved the highest performance, AUC 0.74 (Fig 1B). In this case, adding the SBP PRS did not improve the performance. The most predictive variables in the model (determined using the Shapley interpretability method) were blood pressure, maternal age, and history of preeclampsia in a prior pregnancy (Fig 2).

**Fig 1.**
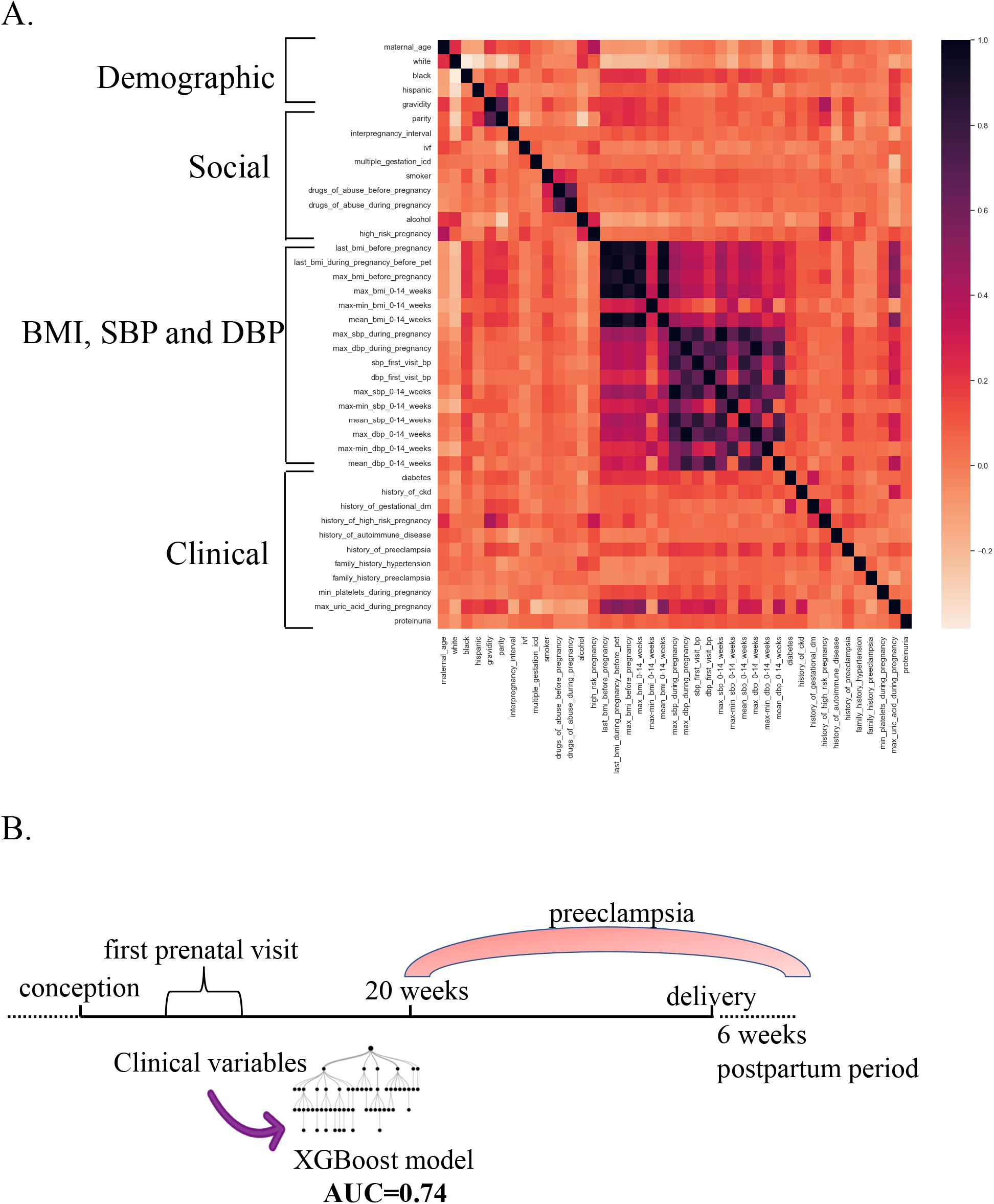
Correlation matrix and preeclampsia predictive model development in early pregnancy, before 14 weeks gestation. Abbreviations: SBP, systolic blood pressure; DBP, diastolic blood pressure, BMI, body mass index

**Fig. 2.**
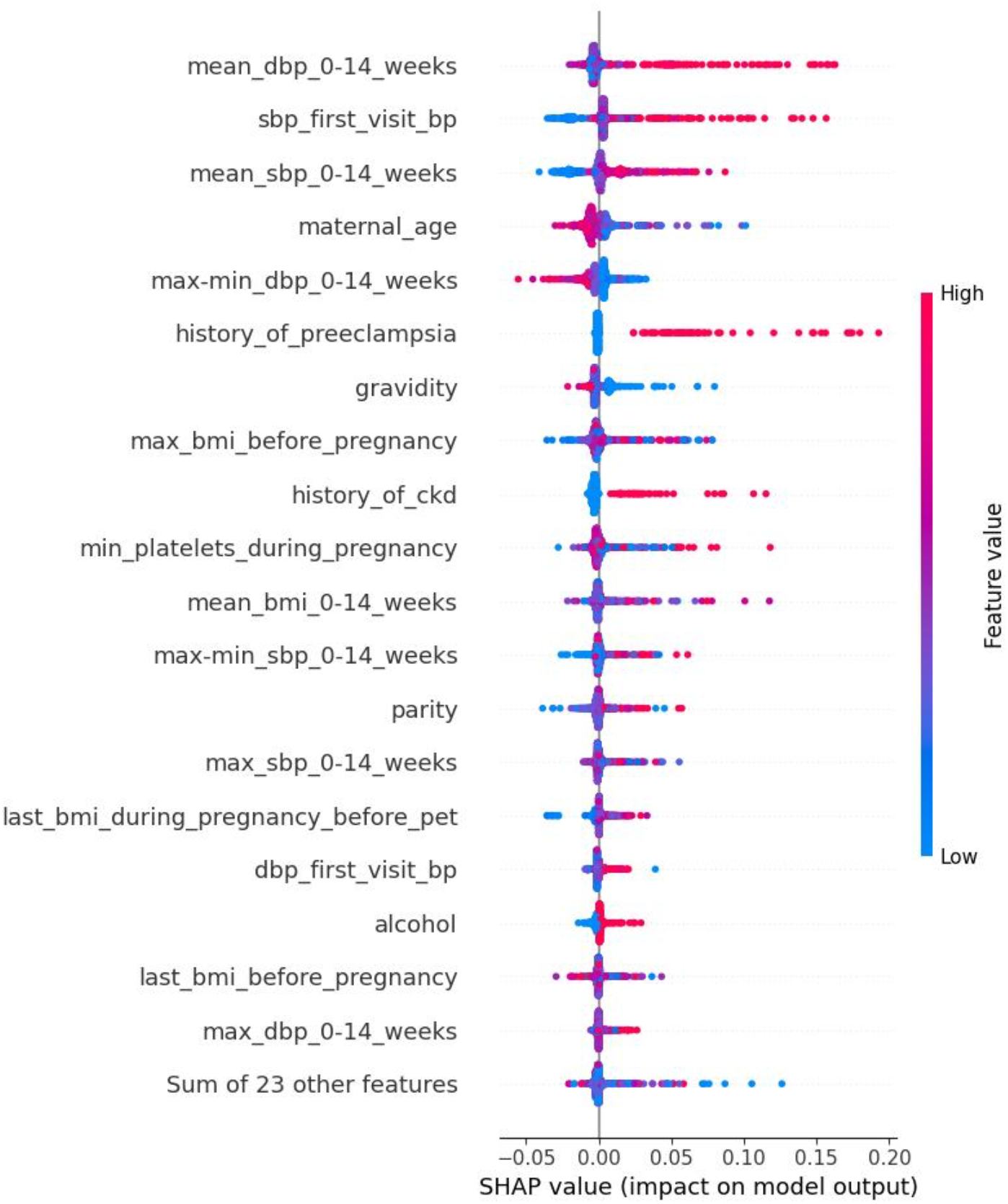
SHapley Additive exPlanations (SHAP) plot of the top variables contributing to the xgboost output in early pregnancy. The horizontal position of each point shows the impact of the feature on the model’s prediction. Red, high feature value; blue low feature value.

By the time of delivery, more clinical information becomes available from scheduled outpatient prenatal visits, which become more frequent during the 3^rd^ trimester (Fig. 3A). The late pregnancy models were generated using clinical information available prior to (but not after) the admission associated with preeclampsia diagnosis (Table 3). In late pregnancy, the linear regression model had an AUC of 0.84 and performed better than the early pregnancy model. Similarly, the machine learning model using clinical risk factors had the best performance of all, AUC 0.91 (Fig. 3B). At this timepoint, the addition of the SBP PRS to the clinical risk factors in the linear regression and machine learning models did not significantly improve the performance. In the best-performing model, the most predictive variables (determined using the Shapley interpretability method) were blood pressure, body mass index, uric acid level, and past medical history of renal disease (Fig. 4).

**Fig 3.**
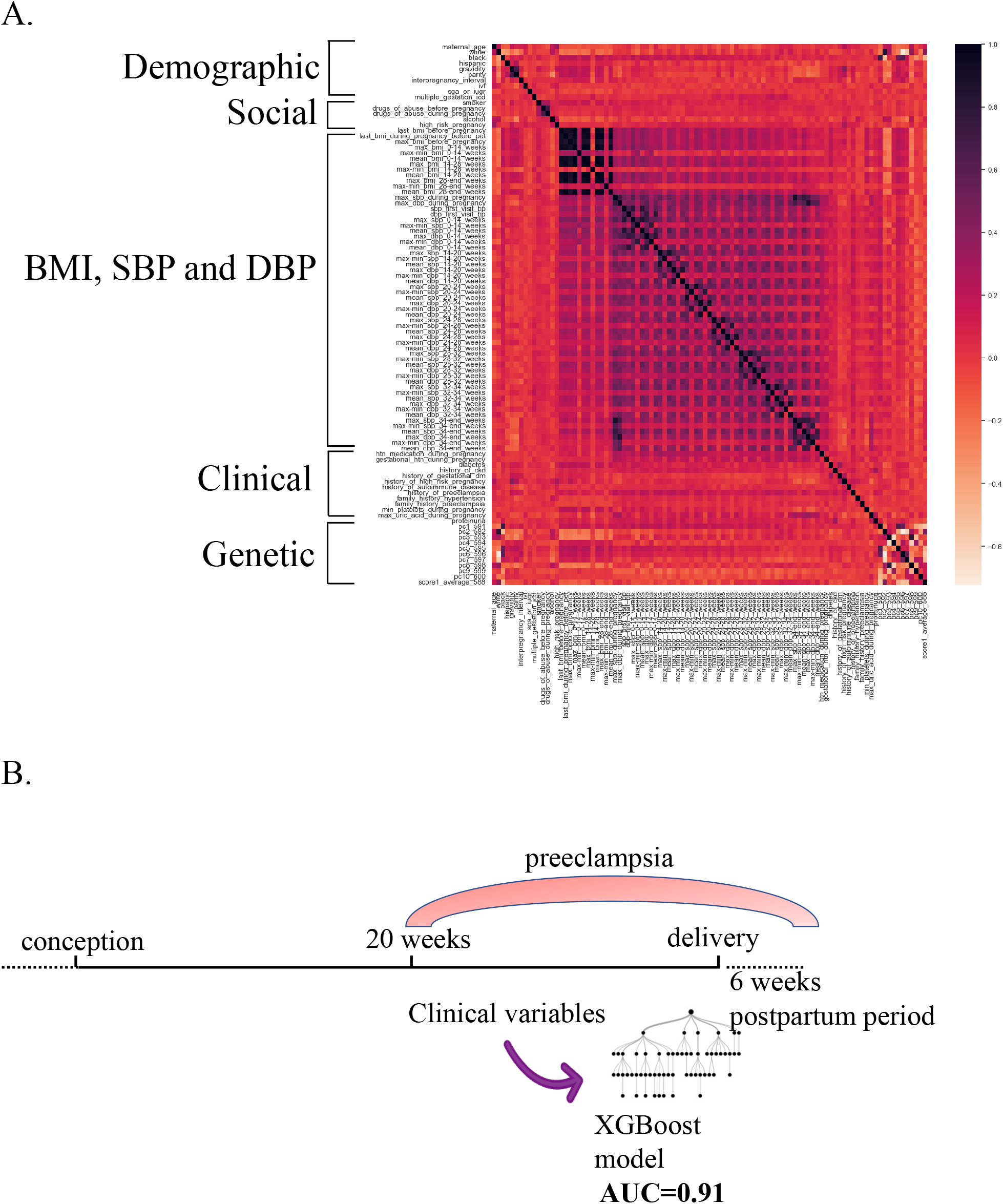
Correlation matrix and preeclampsia predictive model development at late pregnancy, before the admission for delivery. Abbreviations: SBP, systolic blood pressure; DBP, diastolic blood pressure, BMI, body mass index.

**Fig. 4.**
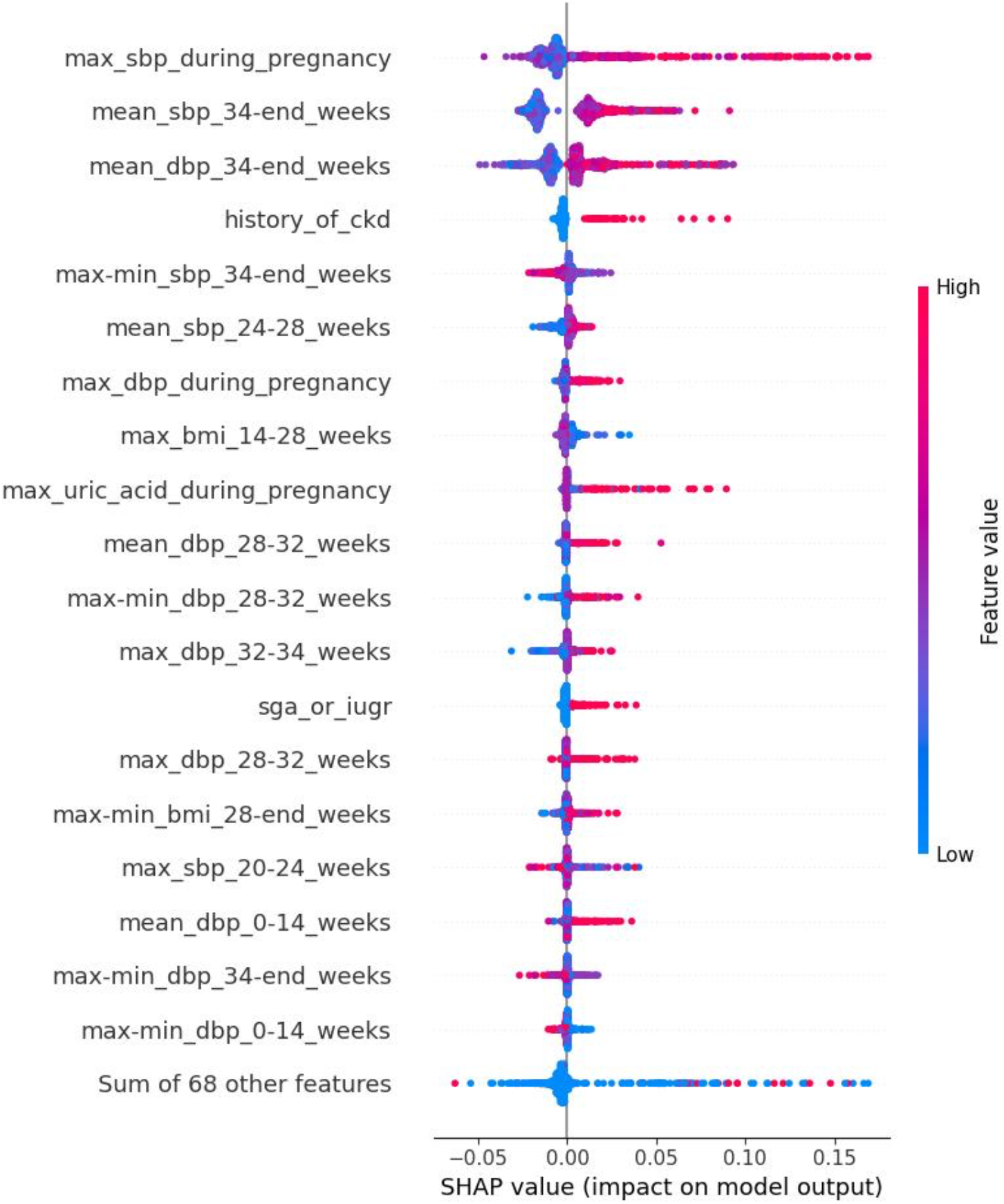
SHapley Additive exPlanations (SHAP) plot of the top variables contributing to the xgboost output in late pregnancy. The horizontal position of each point shows the impact of the feature on the model’s prediction. Red, high feature value; blue low feature value.

## DISCUSSION

Here, we investigated the ability of machine learning and linear regression models based on electronic health records and genetic data to predict preeclampsia. Our results demonstrate that, in a multi-ethnic cohort, SBP PRSs correlate with systolic and diastolic blood pressure during pregnancy, as well as with the diagnoses of gestational and chronic hypertension. In early pregnancy, when less clinical information is available, the addition of SBP PRS to clinical risk factors improves prediction. However, in later pregnancy, when more clinical information is available and overall performance of the predictive models is improved, SBP PRS does not add to the predictive power. In both early and late pregnancy, machine learning models performed better than linear regression models; xgboost in late pregnancy was the most predictive.

In line with prior studies,^29^ we demonstrate that SBP PRS is associated with clinically measured blood pressure and risk of hypertensive disorders. The heritability of hypertensive disorders using PRS is well established, and some recent studies have demonstrated that these findings also translate to hypertensive disorders of pregnancy.^20^ A recent study of preeclampsia and blood pressure PRS has shown a strong disease correlation in Finnish White patients with higher PRS scores.^20^ The maximal blood pressure measured during pregnancy was elevated in the group with the top 25% SBP PRS. We also find that gestational and chronic hypertension, as well as hypertensive disorders of pregnancy (in sensitivity analyses of White patients), are associated with higher PRS scores. We were not able to find a significant relationship between SBP PRS and preeclampsia, hypertensive medication use, and family history in our cohort, likely due to our small sample size. In addition, SBP is only one risk factor for preeclampsia, and future studies using preeclampsia-specific or multi-trait PRS may improve the predictive capacity of polygenic scores. Also, as the current SBP PRS was generated from a White population, future studies utilizing multi-ethnic PRSs, are likely to provide additional insight.

When using only the SBP PRSs and adjusting for the first 10 principal components of ancestry, both linear and machine learning models have low predictive power. The relationships between SBP PRSs and outcomes are non-linear; individuals in the top 2.5% of the SBP PRSs have a disproportionately higher risk of disease and adverse outcomes than those in the lowest 2.5%.^29^ We anticipated that the machine learning approaches, which have the ability to capture complex, nonlinear relationships, will achieve higher predictive power. However, the low overall and inferior performance of the xgboost model is likely due to the small number of variables included in those models and the weak association with the outcome leading to overfitting on the training data.

In early pregnancy, we demonstrate the good predictive power of the linear regression model, which is similar to or better than other studies. ^9,10^ In order to avoid overfitting with a small sample size, we selected only the variables known to be associated with a heightened risk of preeclampsia rather than using all available variables from the electronic medical record; the first approach has previously demonstrated better performance. ^12^ Other models^31,32^ have achieved higher predictive power than ours; however, those included biomarkers like serum placental growth factor and uterine artery pulsatility index, which are not routinely measured in our clinical practice. In addition, we observe increased predictive power with the addition of the genetic risk factors in the early pregnancy linear model. These results highlight the potential value of PRSs to complement the clinical predictions, especially in early pregnancy, when little clinical data is available.

The best-performing model in early pregnancy was xgboost and similarly, others have demonstrated the power of this type of machine learning model in early pregnancy to achieve accurate predictions. ^10,12^ To incorporate information about the rate of change in time-series variables like blood pressure and BMI, we included data routinely recorded at the scheduled office visits. This approach has demonstrated improved predictions. ^10,12^ The early pregnancy screening and prevention of preeclampsia has been associated with improved maternal and neonatal outcomes by 70-89%,^4,5^ and thus, integrating this type of model in clinical practice has the potential for a high-value impact on patient care.

Similar to early pregnancy, the xgboost model in late pregnancy had higher predictive power than the linear regression model, demonstrating the superiority of the machine learning approach. Similar results have been demonstrated by others. ^10^ The strongest predictors for preeclampsia were blood pressure, history of renal disease, and uric acid values, which have been shown by others as well. ^10,12^ Integrating this type of model in clinical practice will aid more accurate personalized prediction and allow for referral of high-risk patients to maternal-fetal medicine specialists and planning for delivery at a tertiary care center.

When including SBP PRSs in the clinical model, we demonstrate similar to others^29^, little to no improvement in the risk prediction. In addition to the considerations in early pregnancy, in late pregnancy, as more clinical information is available, the patients may have expressed the genes that contribute to preeclampsia risk, or the clinical factors may have greater weight relative to the genetic factors. In the future, as better genetic tools and larger datasets become available, this approach may yield improved results.

Our study has several strengths, including detailed data for all patients from multiple visits with a low level of missingness, recent data collected in the past seven years, when the most current clinical guidelines were implemented, ^6^ and data from both tertiary and community hospitals within our large healthcare system.

Our study has several limitations. We had a small cohort of patients; however, accurate predictions using a dataset of similar size have been previously achieved. ^12^ To avoid the risk of overfitting, we limited the types of analyses we performed; for example, we were not able to investigate the predictions of early-onset preeclampsia. In addition, some of the variables are based on billing codes which may be inaccurate and do not reflect disease severity. To overcome this limitation for the preeclampsia phenotype, we developed our own algorithm using the current standard of care and manually validated the cases. We used SBP PRS developed in a White population, which may not have optimally assessed risk in our multiethnic cohort. The SBP PRS we selected were developed from the largest GWAS to date, which was performed in White patients; currently, large multiethnic GWAS are lacking, which is a well-recognized limitation of the field.^33^ Similarly, we were not able to externally validate this model as most large genetic biobanks lack detailed pregnancy information.

## PERSPECTIVES

We demonstrate that models using clinical and genetic data in early and late pregnancy have high predictive power and can accurately predict the individual risk for preeclampsia. In addition, SBP PRSs correlate with risk factors for preeclampsia and improve the predictive power of clinical risk factors in the linear regression model in early pregnancy. Since the machine learning models using clinical data available from routine visits had the highest predictive power, these types of models can be implemented in clinical practice to function within the electronic medical records longitudinally. In this way, the risk predictions can be made available to the treating physician, in addition to the contributing factors, who can advise about prophylactic and therapeutic options, as well as referral to a maternal-fetal medicine specialist. As more pregnancy data in multi-ancestry cohorts becomes available, such strategies can be expanded.

## SUPPLEMENTARY MATERIAL

### Supplementary Methods

#### Limiting Data

Before training the predictive models, we excluded any data that was recorded after the date of delivery for healthy patients, or whichever came first, the preeclampsia diagnosis or delivery date for preeclamptic patients. This data constitutes the dataset for late pregnancy models, while the early pregnancy dataset also excludes any measurements taken after 13 weeks of gestation.

#### Feature Engineering and Selection

To capture trends in vital signs over time, measurements were assessed across different gestational age windows in pregnancy. The systolic and diastolic blood pressures (maximum, difference between maximum and minimum, and mean) were calculated before 14 weeks, 14-20, 20-24, 24-28, 28-32, 32-34, and after 34 weeks. The same measurements were calculated by trimester (<14 weeks, 14-28, and >28 weeks) for BMI since weight and height measurements were recorded less frequently. A simple variance threshold was applied to features prior to cross-validation to remove any features with a variance below the best-performing threshold of 0.04.

#### Cross Validation

We performed a 5-fold cross-validation using sklearn’s StratifiedGroupKFold, which creates folds that tend to preserve the percentage of samples for each class as much as possible, given the constraint of non-overlapping groups between splits. ^34^ For each training and test set created, the data were imputed, scaled, and oversampled. Missing information in binary variables was assumed to be false, while missing continuous variables were imputed with the mean of the feature. Sklearn’s MinMaxScaler was fit on the training data of each fold and modified features to contain values between 0 and 1. We applied several methods to correct for an imbalanced dataset, including SMOTE, BorderlineSMOTE, RandomOverSampler, and RandomUnderSampler. Of these methods, the RandomOversampler produced the best-performing models and was subsequently applied to all cross-validation folds.

#### Regression and XGB Models

Two types of models were trained with the data: a linear regression and an xgboosted ensemble. In addition to the features removed during feature selection, other highly correlated variables were combined or removed when trained on the linear regression. The xgboost ensemble consisted of 3 separate models with a maximum depth of 2, 10, and 20 with 10, 50, or 100 estimators respectively. Higher depth and number of estimators lead to an increased complexity that can be prone to overfitting. Final predictions are the equally weighted average between the individual predictions of each xgboost model. Metrics were calculated on the test set for each fold and averaged together to obtain final statistics.

**Supplemental Table 1.** Variables included in the first prenatal visit (early pregnancy) model.

|  | **Preeclampsia (n=87)** | **No preeclampsia**<br>**(n=1038)**<br>**(includes**<br>**normotensive,**<br>**chronic, and**<br>**gestational HTN)** | **P-value** |
| --- | --- | --- | --- |
| Maternal age, y | 32.9 (29.5 - 36.4) | 33.6 (30.5 - 36.4) | 0.29 |
| Self-reported White race | 60 (69%) | 747 (72%) | 0.75 |
| Self-reported Black race | 13 (15%) | 75 (7%) | 0.013 |
| Self-reported Hispanic ethnicity | 3 (3%) | 45 (4%) | 0.7 |
| Gravidity | 2.0 (1.0 - 3.0) | 2.0 (1.0 - 3.0) | 0.84 |
| Parity | 1.0 (0.25 - 2.0) | 1.0 (1.0 - 2.0) | 0.15 |
| Interpregnancy interval, y | 2.1 (1.6 - 3.2) | 2.3 (1.8 - 3.0) | 0.54 |
| In vitro fertilization | 4 (5%) | 43 (4%) | 0.84 |
| Multiple gestation | 7 (8%) | 39 (4%) | 0.057 |
| Smoking before pregnancy | 21 (24%) | 126 (12%) | < 0.01 |
| Drugs of abuse before pregnancy | 8 (9%) | 59 (6%) | 0.2 |
| Drugs of abuse during pregnancy | 6 (7%) | 46 (4%) | 0.3 |
| Alcohol use before pregnancy | 45 (52%) | 541 (52%) | 0.96 |
| High-risk pregnancy | 51 (59%) | 578 (56%) | 0.72 |
| Maximal BMI before pregnancy, kg/m<sup>2</sup> | 30.8 (25.1 - 36.7) | 26.7 (23.5 - 31.3) | < 0.01 |
| Last BMI before pregnancy, kg/m<sup>2</sup> | 27.7 (23.0 - 33.9) | 24.6 (21.5 - 29.0) | < 0.01 |
| Mean BMI in the period 0-14 gestational weeks, kg/m<sup>2</sup> | 29.7 (23.7 - 34.4) | 24.9 (21.9 - 29.1) | < 0.01 |
| SBP at the first prenatal visit, mmHg | 123.0 (116.0 - 138.0) | 111.0 (104.0 - 120.0) | < 0.01 |
| DBP at the first prenatal visit, mmHg | 78.0 (70.0 - 82.5) | 68.0 (61.0 - 74.0) | < 0.01 |
| History of pregestational diabetes | 8 (9%) | 62 (6%) | 0.25 |
| History of kidney disease before pregnancy | 17 (20%) | 38 (4%) | < 0.01 |
| History of gestational diabetes in a prior pregnancy | 4 (5%) | 54 (5%) | 0.81 |
| History of a prior high-risk pregnancy | 28 (32%) | 334 (32%) | 1 |
| History of autoimmune disease | 7 (8%) | 88 (8%) | 0.89 |
| History of preeclampsia in a prior pregnancy | 23 (26%) | 40 (4%) | < 0.01 |
| Family history of hypertension | 39 (45%) | 464 (45%) | 0.99 |
| Family history of preeclampsia | 1 (1%) | 15 (1%) | 0.82 |
| Minimal platelet count in the period 0-14 gestational weeks | 270 (219 - 315) | 254 (217 - 288) | < 0.01 |
| Maximal uric acid in the period 0-14 gestational weeks, mg/dL | 3.8 (3.3 - 3.4) | 3.0 (2.3 - 3.7) | 0.03 |
| Presence of proteinuria in the period 0-14 gestational weeks | 3 (3%) | 3 (0%) | < 0.01 |
| SBP PRS | 1.8 (1.4 - 2.2) | 1.8 (1.4 - 2.1) | 0.94 |

Median (IQR) for continuous variables; n (%) for categorical variables; p-values for continuous variables based on Kruskal-Wallis rank sum test; for categorical variables based on Fisher's exact or Chi-squared test.
Abbreviations: BMI, body mass index; SBP, systolic blood pressure; DBP, diastolic blood pressure; PCA, principle components of ancestry; PRS, polygenic risk scores.

**Supplemental Table 2.** Variables included in the late pregnancy model (before admission for delivery)

|  | **Preeclampsia (n=87)** | **No preeclampsia (n=1038)**<br>**(includes normotensive, chronic, and gestational HTN)** | **p-value** |
| --- | --- | --- | --- |
| Maternal age, y | 32.3 (29.5 - 36.4) | 33.5 (30.5 - 36.4) | 0.29 |
| Self-reported White race | 60 (69%) | 747 (72%) | 0.75 |
| Self-reported Black race | 13 (15%) | 75 (7%) | 0.013 |
| Self-reported Hispanic ethnicity | 3 (3%) | 45 (4%) | 0.7 |
| Gravidity | 2.0 (1.0 - 3.0) | 2.0 (1.0 - 3.0) | 0.84 |
| Parity | 1.0 (0.25 - 2.0) | 1.0 (1.0 - 2.0) | 0.15 |
| Interpregnancy interval, y | 2.1 (1.6 - 3.2) | 2.3 (1.8 - 3.0) | 0.54 |
| In vitro fertilization | 4 (5%) | 43 (4%) | 0.84 |
| SGA or IUGR | 20 (23%) | 101 (10%) | < 0.01 |
| Multiple gestation | 7 (8%) | 39 (4%) | 0.057 |
| Smoking before pregnancy | 21 (24%) | 127 (12%) | < 0.01 |
| Drugs of abuse before pregnancy | 8 (9%) | 59 (6%) | 0.2 |
| Alcohol use before pregnancy | 46 (53%) | 551 (53%) | 0.98 |
| High-risk pregnancy | 64 (74%) | 715 (69%) | 0.61 |
| Last BMI before pregnancy, kg/m<sup>2</sup> | 27.7 (23.0 - 33.9) | 24.6 (21.5 - 29.0) | < 0.01 |
| Last BMI during pregnancy before preeclampsia diagnosis or delivery, kg/m<sup>2</sup> | 32.9 (28.6 - 37.9) | 30.0 (26.4 - 33.6) | < 0.01 |
| Maximal BMI before pregnancy, kg/m<sup>2</sup> | 30.8 (25.1 - 36.7) | 26.7 (23.5 - 31.3) | < 0.01 |
| Maximal SBP during pregnancy, mmHg | 151.0 (142.0 - 160.5) | 129.0 (120.0 - 138.0) | < 0.01 |
| Maximal DBP during pregnancy, mmHg | 93.0 (87.2 - 98.7) | 80.0 (74.0 - 85.7) | < 0.01 |
| SBP at the first prenatal visit, mmHg | 122.0 (115.5 - 136.0) | 111.0 (104.0 - 120.0) | < 0.01 |
| DBP at the first prenatal visit, mmHg | 75.0 (69.0 - 81.0) | 68.0 (60.0 - 74.0) | < 0.01 |
| Prescription of antihypertensive medication during pregnancy | 11 (13%) | 5 (0%) | < 0.01 |
| Diagnosis of gestational hypertension during pregnancy | 17 (20%) | 36 (3%) | < 0.01 |
| History of pregestational diabetes | 8 (9%) | 62 (6%) | 0.25 |
| History of kidney disease before pregnancy | 17 (20%) | 38 (4%) | < 0.01 |
| History of gestational diabetes in a prior pregnancy | 4 (5%) | 54 (5%) | 0.81 |
| History of a prior high-risk pregnancy | 28 (32%) | 334 (32%) | 1 |
| History of autoimmune disease | 7 (8%) | 88 (8%) | 0.89 |
| History of preeclampsia in a prior pregnancy | 23 (26%) | 40 (4%) | < 0.01 |
| Family history of hypertension | 39 (45%) | 464 (45%) | 0.99 |
| Family history of preeclampsia | 1 (1%) | 15 (1%) | 0.82 |
| Minimal platelet count in pregnancy before preeclampsia diagnosis or delivery | 212 (168 - 270) | 212 (181 - 248) | 0.19 |
| Maximal uric acid in pregnancy before preeclampsia diagnosis or delivery, mg/dL | 5.0 (4.1 - 5.8) | 4.3 (3.5 - 5.2) | < 0.01 |
| Proteinuria in pregnancy before preeclampsia diagnosis or delivery | 12 (14%) | 7 (1%) | < 0.01 |
| SBP PRS | 1.8 (1.4 - 2.2) | 1.8 (1.4 - 2.1) | 0.94 |

Median (IQR) for continuous variables; n (%) for categorical variables; p-values for continuous variables based on Kruskal-Wallis rank sum test; for categorical variables based on Fisher's exact or Chi-squared test.
Abbreviations: BMI, body mass index, SBP, systolic blood pressure, SGA, small for gestational age, IUGR, intrauterine growth retardation, DBP, diastolic blood pressure, PCA, principle components of ancestry, PRS, polygenic risk scores.

**Suppl. Fig. 1.**
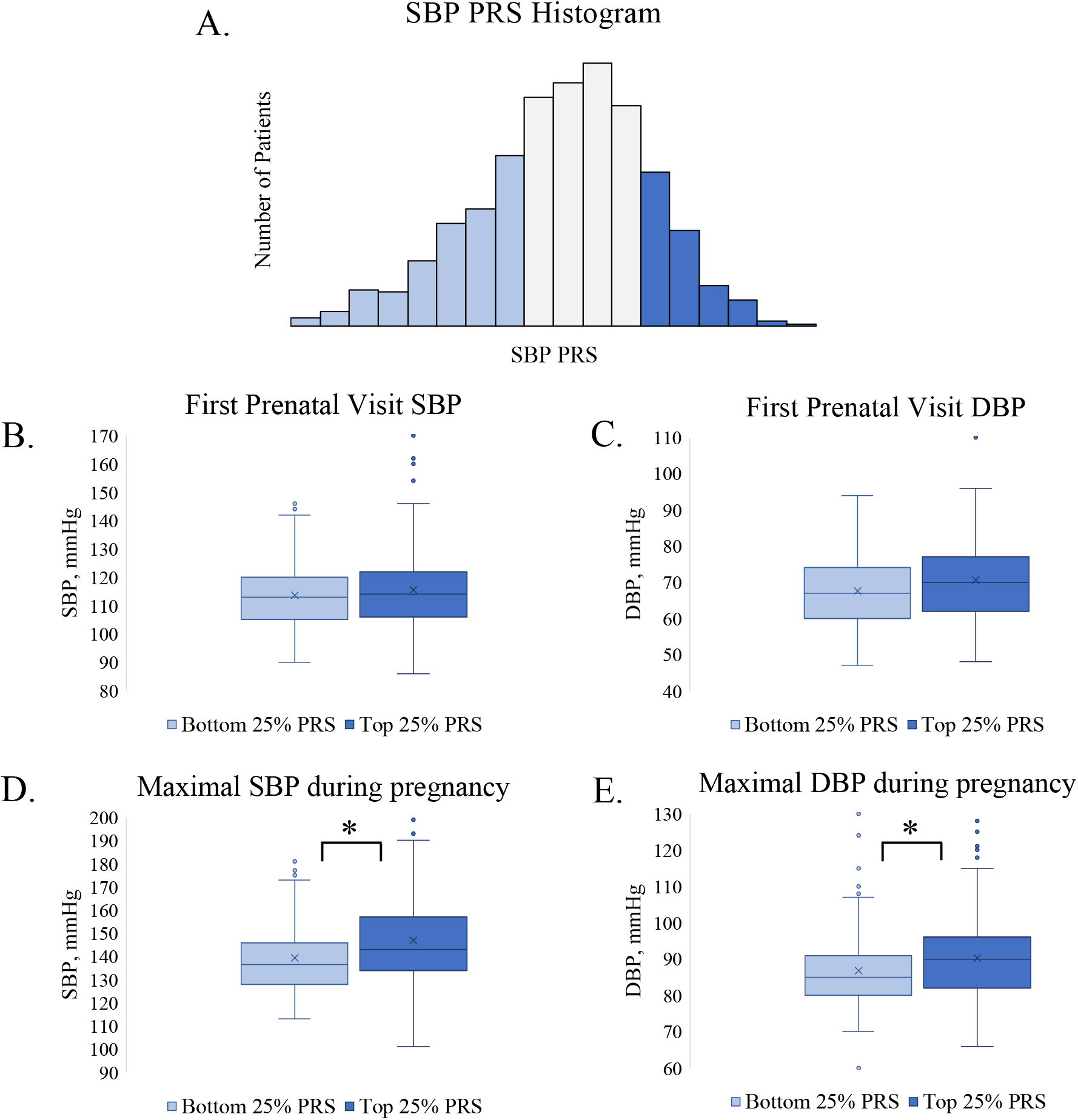
Relationship between BP and SBP PRS in patients with self-reported White race. A. PRS histogram, B. SBP at the first prenatal visit, C. DBP at the first prenatal visit, D. Max SBP during pregnancy E. Max DBP during pregnancy * p<0.05 Abbreviations: SBP, systolic blood pressure; DBP, diastolic blood pressure; PRS polygenic risk score

**Suppl. Fig. 2.**
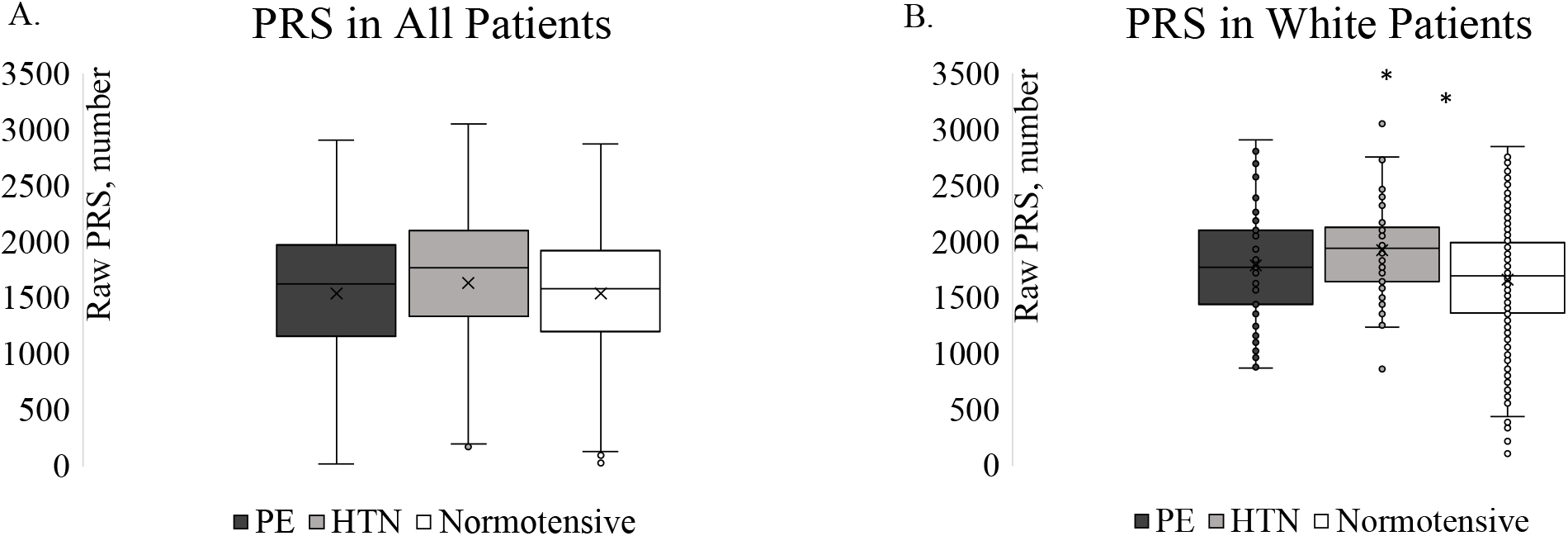
Polygenic risk scores (PRS) in patients with preeclampsia (PE), chronic and gestational hypertension (HTN), and normotension.(A) All patients, (B) Patients with self-reported White race.

**Suppl. Fig 3A.**
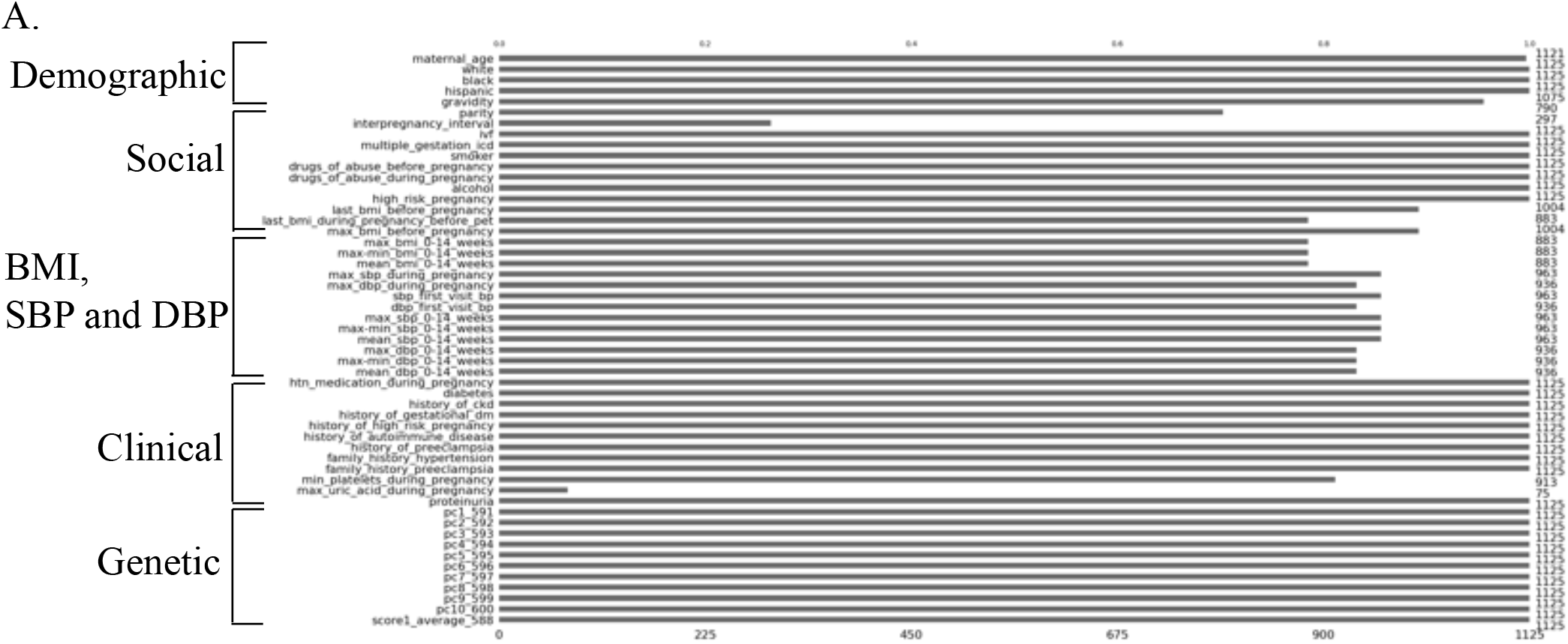
Missing data or data not collected in early pregnancy. Available data is shown in black.

**Suppl. Fig 3B.**
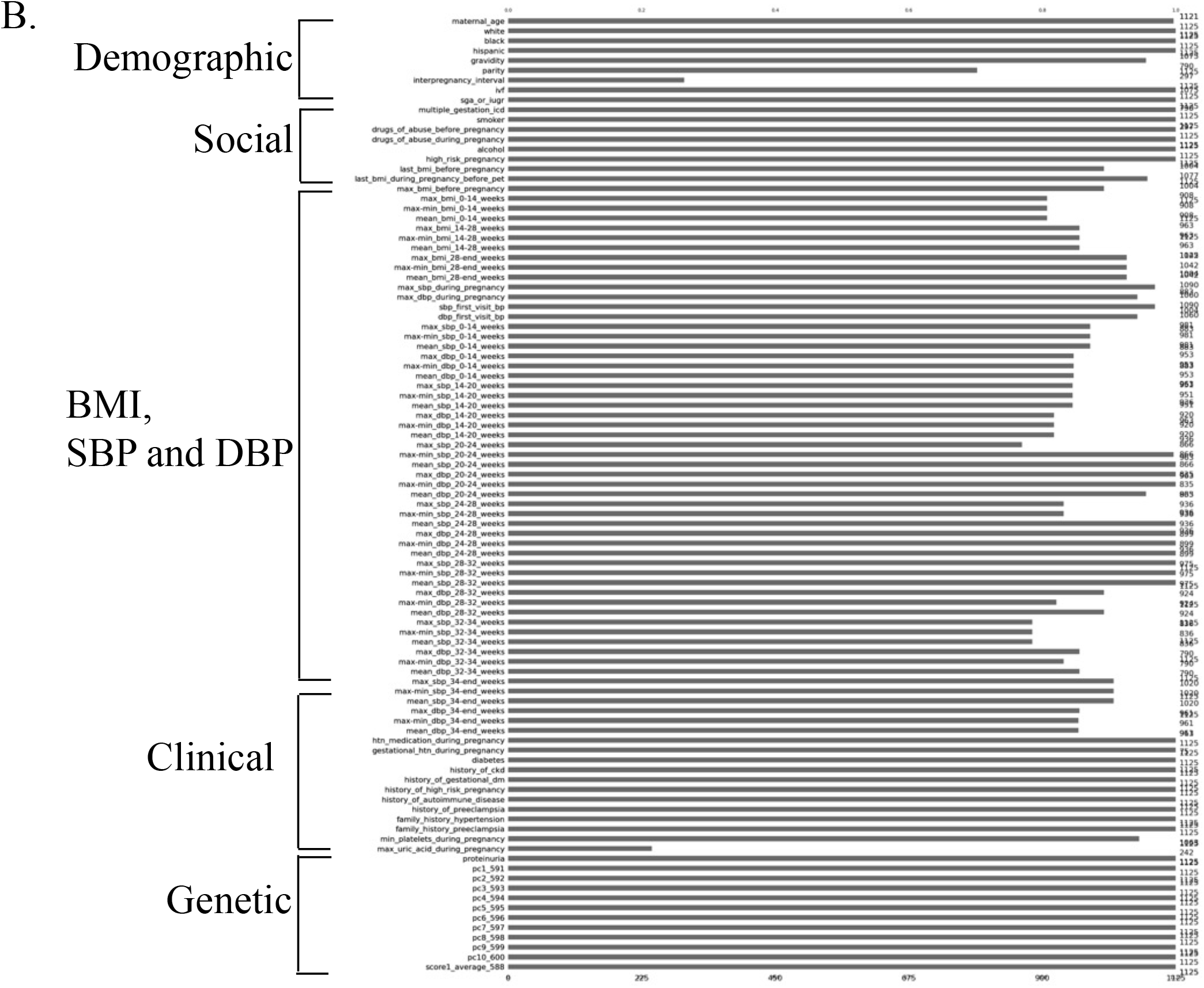
Missing data or data not collected in late pregnancy. Available data is shown in black.

## NON-STANDARD ABBREVIATIONS AND ACRONYMS

AUC: area under the receiver operator curve
BMI: body mass index
DBP: diastolic blood pressure
GWAS: genome-wide association study
IUGR: intrauterine growth restriction
PCA: principle components of ancestry
PRS: polygenic risk score
SBP: systolic blood pressure
SGA: small for gestational age
SNP: single-nucleotide polymorphism
XGB: xgboost

